# Health Utility Value of Overactive Bladder in Japanese Older Adults

**DOI:** 10.1101/2024.07.31.24311331

**Authors:** Takashi Yoshioka, Kenji Omae, Satoshi Funada, Tetsuji Minami, Rei Goto

## Abstract

**Objectives:** To determine the health utility values (HUVs) of overactive bladder (OAB) among adults aged ≥65 years and to assess the HUV decrements (disutilities) of OAB according to its severity.

**Methods:** This cross-sectional Internet-based study was conducted between 2 and 9 November 2023, with quota sampling with equal probability for each gender and age group (age 65–74 years and ≥75 years). OAB was defined as an urgency score of ≥2 points and a total score of ≥3 points based on the Overactive Bladder Symptom Score. OAB severity was categorized as mild (total score, ≤5 points) or moderate-to-severe (total score, 6–15 points). HUVs were measured using the EuroQol 5-Dimension 5-Level value set for the Japanese population. Multivariable linear regression models were fitted to estimate the covariate-adjusted disutilities of OAB, with eight covariates selected based on previous studies.

**Results:** Among the 998 participants (51.9% male; mean age, 73.2 years), 158 (15.9%) had OAB, of whom 87 (8.8%) had moderate-to-severe OAB. The mean HUVs for participants with mild and moderate-to-severe OAB were 0.874 and 0.840, respectively, which were lower compared with the HUV for those without OAB (0.913). After adjusting for relevant covariates, disutilities (95% confidence intervals [CIs]) for mild and moderate-to-severe OAB were –0.0334 (–0.0602 to –0.0066) and –0.0591 (–0.0844 to –0.0339), respectively.

**Conclusions:** This study examined HUVs in older Japanese adults with and without OAB. The results demonstrate that increased OAB severity is associated with greater disutility.

## Introduction

Overactive bladder (OAB), characterized by urinary urgency with or without urgent urinary incontinence, is associated with a substantial disease burden and significant economic impact [1]. The extensive disease burden of OAB includes a decrease in health-related quality of life (HRQoL), mood disorders, and various comorbid or consequential conditions (*e.g.* constipation, fecal incontinence, and irritable bowel syndrome) [2]. The prevalence of OAB increases with age [3]. Furthermore, evidence has demonstrated a longitudinal association between OAB or its treatment with anticholinergics and an increased risk of falls, which are the most critical outcomes in older adults [4]. Consequently, patients with OAB have higher healthcare resource utilization than those without the condition, which contributes to a substantial economic burden [5]. The high global prevalence of OAB, estimated at 15.5% between 2007 and 2020, and its high healthcare cost of $82.6 billion by 2020 in the United States [1, 6] have drawn significant attention from both policymakers and clinicians worldwide.

OAB is widely managed in various healthcare settings, including primary care, urology, and urogynecology [7]. Treatment options for OAB include behavioral therapy and pharmacotherapy, such as antimuscarinic and β-adrenergic agents [7]. Surgical treatments are available for refractory cases, including intradetrusor injection of onabotulinum toxin A, sacral neuromodulation, and percutaneous tibial nerve stimulation [7]. Although these therapeutic options have the potential to relieve patients’ symptoms and improve their HRQoL, they also raise concerns about the increasing economic burden on healthcare systems owing to higher treatment costs. Consequently, the importance of effectively allocating finite healthcare resources for the management of OAB is becoming increasingly recognized. In fact, the National Institute for Health and Care Excellence (NICE), an independent organization providing evidence-based guidance to inform healthcare decisions in the United Kingdom (UK), is currently conducting a health economic evaluation (HEE) of vibegron [8], a novel β3-adrenergic agonist with fewer antimuscarinic side effects compared with traditional OAB medications [9]. Considering the example of the NICE in the UK, new medical technologies for OAB, such as vibegrons, may also be subject to HEEs in other countries, such as Japan.

To make HEEs useful for policy decision-making, it is essential to assess their clinical effectiveness (*e.g.* quality-adjusted life years [QALYs]) using generic measures that can be collected for any disease. QALYs are calculated based on health utility values (HUVs) measured using generic preference-based measures (PBMs) rather than disease-specific ones [10]. Importantly, according to the NICE technology appraisal guidance, HUVs should be calculated using the EuroQoL 5-dimensions (EQ-5D), with data collected directly from patients in the country where the evaluation is being conducted (in the case of NICE, the UK) [11]. Previous studies have evaluated the association between OAB symptoms and HUVs using EQ-5D scores in the United States, United Kingdom, Spain, and South Korea [12–14]. However, these studies have utilized the older version of the EQ-5D, the three-level EQ-5D version (EQ-5D-3L), which raises concerns about the potential overestimation of positive effects and underestimation of negative effects [15]. The newer version of the EQ-5D, the five-level EQ-5D version (EQ-5D-5L), was developed to overcome the ceiling effects and low sensitivity inherent in EQ-5D-3L [16]. Ideally, the estimation of HUVs based on EQ-5D-5L is essential. However, few studies have used this instrument.

To address the unmet needs of global policies regarding HEEs of OAB, this study aimed to describe the HUVs of patients with OAB and assess the impacts of OAB on HUVs using EQ-5D-5L among older adults, the most vulnerable and policy-relevant subgroup, using an Internet-based survey.

## Participants and methods

### Study design and participants

This cross-sectional study was conducted as part of a research project aimed at developing an EQ-5D-5L value set for the Japanese general older adults [16]. To obtain responses from a sample of older adults, we used a sampling method that ensured a 1:1 ratio for both sex and age groups (age 65–74 and ≥75 years). The target sample size was 1,000 participants. The sampling strategy and sample size were determined based on a previous study [17]. All participants were recruited from a survey panel provided by a Japanese Internet research agency (Intage, Inc., https://www.intage.co.jp/english/) with approximately 2.6 million panelists and their demographic information. The survey was conducted between 2 and 9 November 2023. We designed the survey to prevent participants from proceeding to the next question without providing a valid response, thereby eliminating missing data. The survey questionnaire and response options are presented in **Method S1**.

### Inclusion and exclusion criteria

All respondents were included in the analysis. However, participants who responded ‘prefer not to answer’ to at least one of the questionnaires except for income items were excluded from this study.

### Exposures (overactive bladder)

The variable of interest in this study was the presence of OAB. OAB was assessed using the Overactive Bladder Symptom Score (OABSS), a validated questionnaire developed in Japanese [18]. The OABSS is a self-reported questionnaire consisting of four domains (daytime frequency, 0–2 points; nighttime frequency, 0–3 points; urgency, 0–5 points; urgency incontinence, 0–5 points), with a total score of 15 points [18] A higher score indicates more severe OAB symptoms. Based on previous studies and decision modelling on HEEs, OAB was defined as an urgency score of ≥2 points and a total score of ≥3 points, and the severity of OAB was categorized as mild (OAB with a total score of ≤5 points) or moderate-to-severe (OAB with a total score of 6–15 points) [14].

### Outcomes (health utility values)

Our primary outcome of interest was HUVs measured using the EQ-5D-5L instrument. HUVs are an indicator of an individual’s current health state, where 0 represents death or the worst imaginable health state and 1 represents perfect health, with higher scores indicating better health status [10]. In HEEs, HUVs are calculated using PBMs developed with choice methods (*e.g.* standard gambling or time trade-off) as reference standards [10, 11]. The EQ-5D-5L is a PBM developed by the EuroQoL Group that is used to measure HUVs based on time trade-off methods [10, 11]. Importantly, the EQ-5D-5L has been translated and validated in Japanese, demonstrating substantial reliability and validity for calculating HUVs in the general Japanese population [17].

In this study, we used the EuroQoL Visual Analogue Scale (EQ VAS) score as another measure of health utility. The EQ VAS is a scale developed by the EuroQoL Group that measures the current health state using a VAS on a vertically oriented ruler with markings from 0 to 100 [17]. On the EQ-VAS, 0 represents death or the worst imaginable health state, and 100 indicates perfect health. The EQ VAS has often been used in conjunction with the EQ-5D in similar studies [14], and consistent results have been reported. Therefore, we used the EQ-VAS score as an outcome measure for sensitivity analysis in this study.

This study used the online formats of the EQ-5D-5L and EQ VAS. The research protocol was pre-registered on the EuroQoL website [16], and permission was obtained to use these instruments. The online versions were administered in strict accordance with the manufacturers’ instructions. The EuroQoL registration number for this study is 59331.

### Covariates

The covariates of interest in this study were demographic, socioeconomic, and health-related variables. These variables were selected from previous studies on OAB that focused on the factors that could potentially influence HUVs. The demographic factors included age (continuous), gender (male or female), and body mass index BMI (continuous). Socioeconomic status included educational attainment (high school education or less and college education or more) and income [6]. As a proxy for income, this study used equivalent household income, derived by adjusting total household income by the square root of household size (then grouped by median, with ranges provided: lower half, JPY 0–2.8 million; upper half, JPY ≥2.9 million), along with a declined to answer category. Health-related factors included smoking status (never/past/current), alcohol consumption (never/past/current), and self-reported coexisting physical and psychiatric comorbidities. Comorbidity data were used to record the presence or absence of the following diseases: hypertension, dyslipidemia, diabetes, low back pain, ischemic heart disease, stroke, chronic kidney disease, stress urinary incontinence, depression, and other psychiatric disorders [5, 19–22].

### Statistical analyses

This study used three analytical approaches. First, a primary analysis was performed targeting all respondents. Second, subgroup analyses were performed, stratified based on sex (male/female) and age (65–74 years old/≥75 years), assuming that the impacts of OAB on HUVs may vary according to these factors. Third, an exploratory analysis was performed to evaluate the impact of each of the four domains constituting the OAB on HUVs. In all analyses, we initially described the baseline characteristics, including the prevalence of OAB and HUVs. Subsequently, we described the mean (standard deviation [SD]) and median (interquartile range [IQR]) of HUVs based on summary statistics for each group for all analyses. Finally, we estimated the adjusted HUV decrements (disutilities) using a multivariable linear regression analysis after adjusting for covariates. In the multivariable linear regression analysis with ordinal categories (non-OAB/mild OAB/moderate-to-severe OAB), a linear trend test was conducted to assess linearity. To confirm the consistency of the results, a sensitivity analysis was performed in the primary analysis to fit a similar linear regression model using the EQ VAS score as the outcome measure.

For all regression models, β coefficients, 95% confidence intervals (CIs), and *p*-values were calculated. Statistical significance was set at *p*<0.05. All analyses were performed using STATA version 18.0 (Stata Corp, College Station, Texas, USA).

### Ethical considerations

All respondents completed online questionnaires after providing digital informed consent indicating their willingness to participate in the survey. Participants received monetary points as an incentive for participation. All the procedures adhered to the ethical guidelines of the 1975 Declaration of Helsinki and its subsequent 2013 revision. This study was approved by the Institutional Review Board of Keio University School of Medicine (approval number: 20221120). This study followed the Strengthening the Reporting of Observational Studies in Epidemiology (STROBE) guidelines for cross-sectional studies.

## Results

### Baseline characteristics

Of the 1,099 respondents, 994 (90.7%) were included in the analysis (**Figure 1**). The median age of the participants was 73.2 (SD, 5.3) years, and 516 (51.9%) were male. Regarding socioeconomic indicators, 557 (56.0%) participants had college or higher education. Regarding health-related factors, 107 (10.8%) participants were current tobacco users, 519 (52.2%) were current alcohol users, 486 (48.9%) had hypertension, 166 (16.7%) had diabetes, 114 (11.5%) had stress urinary incontinence, and 50 (5.0%) had depression. The mean OABSS was 2.7 (SD, 2.2). The mean HUV was 0.90 (SD, 0.11), and the mean EQ VAS score was 75.2 (SD, 17.6) (**Table 1**).

**Figure 1.**
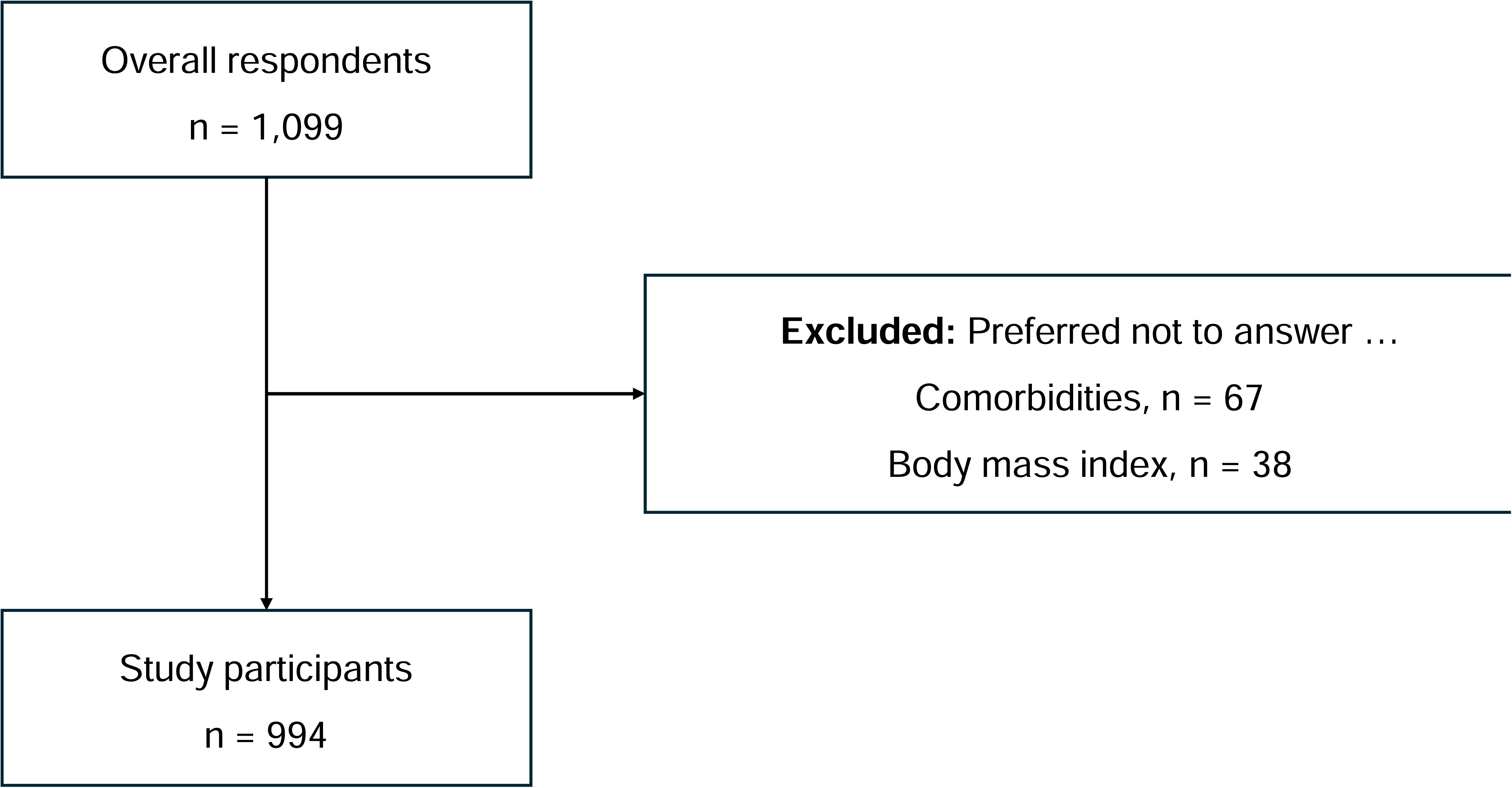
Flow diagram of this study

**Table 1.**
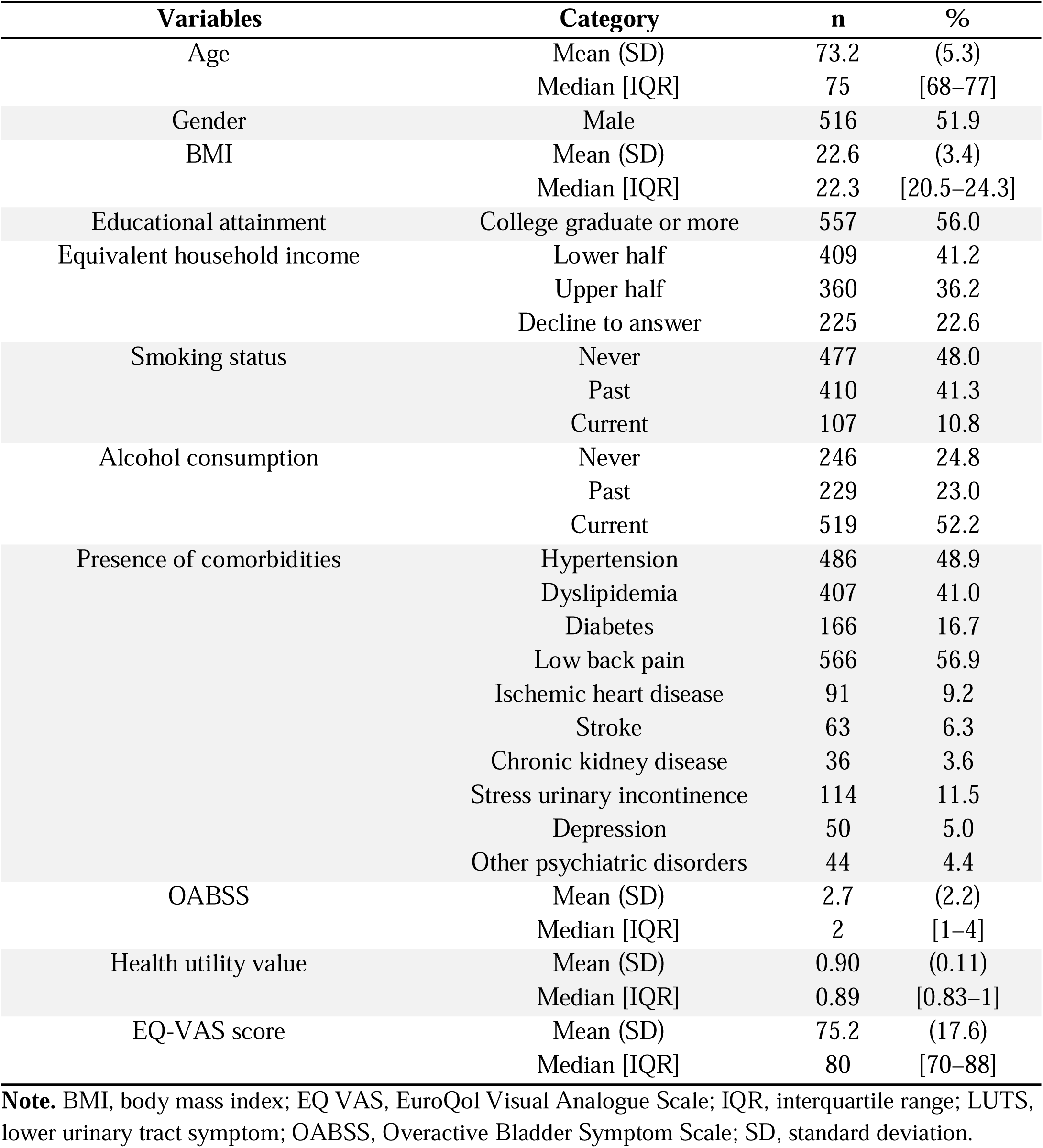
Baseline characteristics of respondents (n=994)

### Health utility values based on overactive bladder status

**Table 2** presents a summary of the HUVs stratified according to the presence and severity of OAB. The mean HUV for participants without OAB was 0.913 (SD, 0.105), whereas that for those with OAB was 0.855 (SD, 0.145). When categorizing OAB severity into mild and moderate-to-severe OAB, the mean HUV for participants with mild OAB was 0.874 (SD, 0.131), and for those with moderate-to-severe OAB, it was 0.840 (SD, 0.155).

**Table 2.** Prevalence of OAB and utility values among overall respondents.

| OAB severity<sup>*</sup> | n (%) | Mean utility value (SD) | Median utility value [IQR] |
| --- | --- | --- | --- |
| **Non-OAB** | **836 (84.1)** | **0.913 (0.105)** | **0.894 [0.867–1]** |
| **Any OAB** | **158 (15.9)** | **0.855 (0.145)** | **0.885 [0.784–1]** |
| Mild OAB | 71 (7.1) | 0.874 (0.131) | 0.895 [0.823–1] |
| Moderate-to-severe OAB | 87 (8.8) | 0.840 (0.155) | 0.844 [0.759–1] |

**Note.** IQR, interquartile range; OAB, overactive bladder; SD, standard deviation.
<sup>\*</sup>Mild OAB is defined as domain 3 (urgency) of overactive bladder symptom score (OABSS) $\geq 2$ and total score of $3 \leq \text{OABSS} \leq 5$ , and moderate-to-severe OAB is defined as domain 3 of OABSS $\geq 2$ and total score of $6 \leq \text{OABSS} \leq 15$ .

### Disutilities of overall respondents

**Table 3** presents the results of the multivariable linear regression analyses. After adjusting for covariates, the estimated disutility for the presence of OAB was –0.0472 (95% CI, –0.0664 to –0.0279; *p*<0.001). The estimated disutilities for mild and moderate-to-severe OAB were –0.0334 (95% CI, –0.0602 to –0.0066; *p*=0.014) and –0.0591 (95% CI, –0.0844 to –0.0339; *p*<0.001), respectively, indicating a linear relationship (*p* for trend<0.001). The sensitivity analysis using the EQ VAS as the outcome variable showed similar results (presence of OAB, β=–5.42, 95% CI –8.47 to –2.37, *p*=0.001; mild OAB, β=–4.36, 95% CI –8.60 to –0.12, *p*=0.044; moderate-to-severe OAB, β=–6.26, 95% CI –10.26 to –2.27, *p*=0.002; *p* for trend<0.001).

**Table 3.** Results of multivariable linear regression models in the main and sensitivity analyses.

| Variables | Category | $\beta^*$ | SE | 95% CI | *p*-value | *p* for trend |
| --- | --- | --- | --- | --- | --- | --- |
| Main analysis (outcome measure: EQ-5D-5L) |  |  |  |  |  |  |
| Presence or absence of OAB | Non-OAB |  |  | Reference |  | Not applicable |
|  | Any OAB | −0.0472 | 0.0098 | −0.0664 to −0.0279 | <0.001 |  |
| Based on OAB severity | Non-OAB |  |  | Reference |  | <0.001 |
|  | Mild OAB | −0.0334 | 0.0136 | −0.0602 to −0.0066 | 0.014 |  |
|  | Moderate-to-severe OAB | −0.0591 | 0.0129 | −0.0844 to −0.0339 | <0.001 |  |
| Sensitivity analysis (outcome measure: EQ-VAS) |  |  |  |  |  |  |
| Presence or absence of OAB | Non-OAB |  |  | Reference |  | Not applicable |
|  | Any OAB | −5.42 | 1.55 | −8.47 to −2.37 | 0.001 |  |
| Based on OAB severity | Non-OAB |  |  | Reference |  | <0.001 |
|  | Mild OAB | −4.36 | 2.16 | −8.60 to −0.12 | 0.044 |  |
|  | Moderate-to-severe OAB | −6.26 | 2.04 | −10.26 to −2.27 | 0.002 |  |

**Note.** EQ-5D-5L, the 5-level EuroQoL 5 dimensions version; EQ VAS, EuroQol Visual Analogue Scale; CI, confidence interval; OAB, overactive bladder; SE, standard error.
\* Adjusted for age, sex, body mass index, educational attainment, equivalent household income, smoking status, alcohol consumption, and presence of comorbidities (hypertension, dyslipidemia, diabetes, low back pain, ischemic heart disease, stroke, chronic kidney disease, stress urinary incontinence, depression, and other psychiatric disorders).

### Results of the subgroup analysis

**Tables S1** and **S2** present the baseline characteristics of the sex and age subgroups, respectively. **Table S3** summarizes the HUVs for sex and age subgroups of HUVs. In the sex subgroup, regardless of the presence of OAB, females (n=478) had lower mean HUVs than males (n=516) (*e.g.* presence of any OAB: male, 0.872 [SD, 0.128]; female, 0.821 [SD, 0.173]). In the age subgroup, regardless of the presence of OAB, the 75–94-year-old group (n=502) showed lower HUVs compared with the 65–74-year-old group (n=492) (*e.g.* presence of any OAB: 65–74 years old, 0.889 [SD, 0.130]; 75–94 years old, 0.829 [SD, 0.152]).

The results of the multivariable linear regression analysis are shown in **Figure 2**. In the gender subgroup, a substantial disutility was observed in females with moderate-to-severe OAB (β=–0.1026; 95% CI, –0.1510 to –0.0542; *p*<0.001). In the age subgroup, substantial disutilities were observed in the 75–94-year-old group with mild OAB (β=–0.0688; 95% CI, –0.1106 to –0.0271; *p*=0.001) and moderate-to-severe OAB (β=–0.0664; 95% CI, –0.1006 to –0.0322; *p*<0.001).

**Figure 2.**
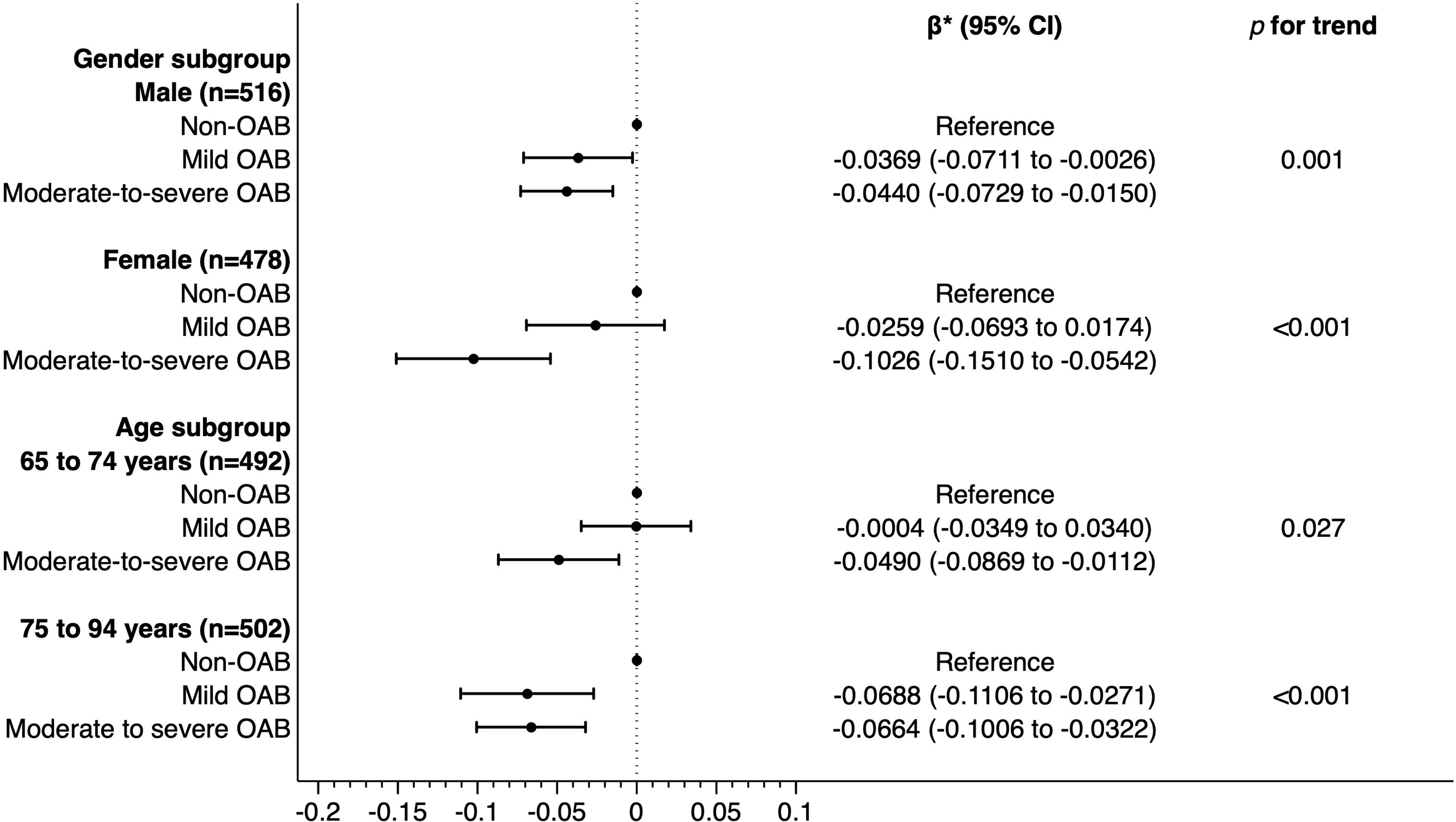
Results of multivariable linear regression models in the subgroup analyses *Adjusted for age, gender, body mass index, educational attainment, equivalent household income, smoking status, alcohol consumption, and presence of comorbidities (hypertension, dyslipidemia, diabetes, low back pain, ischemic heart disease, stroke, chronic kidney disease, stress urinary incontinence, depression, and other psychiatric disorders). **Abbreviations:** CI, confidence interval; OAB, overactive bladder.

### Results of the exploratory analysis

**Table S4** presents a summary of the HUVs for each domain and the score of the OABSS. The higher OABSS scores indicated the lower mean HUVs in the three domains, excluding urgency urinary incontinence. The results of the multivariable linear regression analysis after rounding scores of <2.5% of the total are shown in **Figure 3**. After adjusting for covariates, disutilities were observed for each score in the three domains, excluding daytime frequency (*e.g.* nighttime frequency: 1 point, β=–0.0089, 95% CI –0.0276 to 0.0099, *p*=0.035; 2 points, β=–0.0193, 95% CI –0.0411 to 0.0025, *p*=0.082; 3 points, β=–0.0409, 95% CI –0.0682 to –0.0136, *p*=0.003). A linear relationship between scores and disutilities was observed in all domains, except for daytime frequency (*p* for trend: nighttime frequency, 0.003; urgency, <0.001; and urgency incontinence, <0.001).

**Figure 3.**
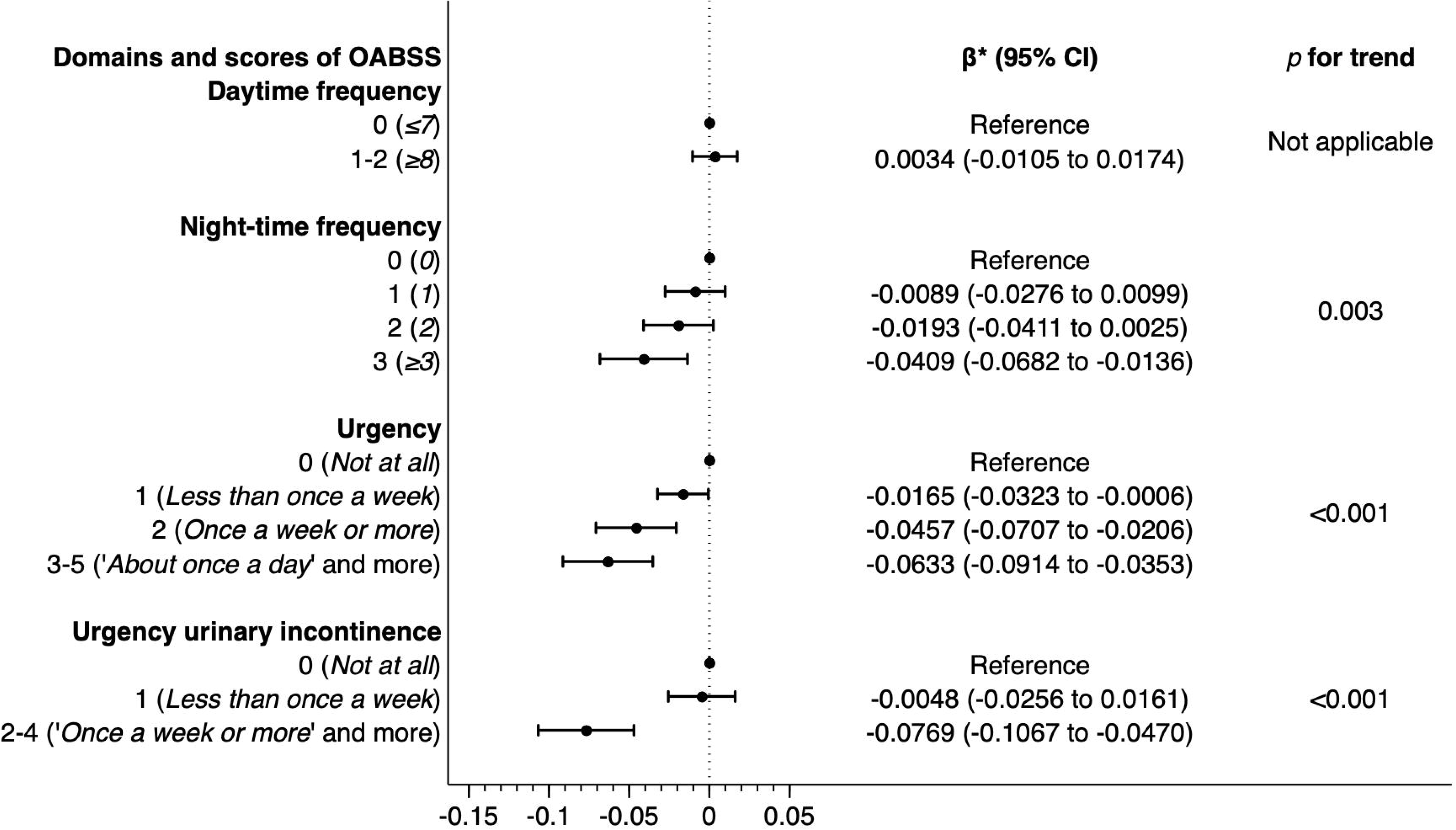
Results of multivariable linear regression models in the exploratory analyses *Adjusted for age, gender, body mass index, educational attainment, equivalent household income, smoking status, alcohol consumption, and presence of comorbidities (hypertension, dyslipidemia, diabetes, low back pain, ischemic heart disease, stroke, chronic kidney disease, stress urinary incontinence, depression, and other psychiatric disorders). **Abbreviations:** CI, confidence interval; OABSS, overactive bladder symptom score.

## Discussion

This study described the HUVs of Japanese adults aged ≥65 years with or without OAB and estimated the associated disutility of OAB using regression models. The results demonstrated that the mean HUVs of individuals with OAB were lower and the disutility of OAB was greater, depending on the severity of the condition. Subgroup analyses suggested that females and those aged 75 years might experience greater disutility owing to OAB than their counterparts. Furthermore, exploratory analyses indicated the possibility of score-proportional disutility in all domains of the OABSS, except daytime frequency.

Previous studies have evaluated the associations of OAB with HUVs in several countries. Kay *et al*. conducted a study estimating EQ-5D-3L-based HUVs from the Incontinence-Specific Quality of Life Questionnaire in 2,505 patients with idiopathic or neurogenic OAB in the United States and Europe [12]. Similarly, Ruiz *et al*. used the Overactive Bladder Symptom and Health-Related Quality of Life Questionnaire in 246 patients with OAB to estimate the effects of OAB symptoms on EQ-5D-3L-based HUVs in Spain [13]. Additionally, Kim *et al*. assessed the effects of OAB severity as defined by the OABSS on EQ-5D-3L-based HUVs in 226,867 participants (including 12,303 patients with OAB) from the 2012 Korean Community Health Survey [14]. These studies shared two common features. First, they estimated the associations of OAB with HUVs using OAB measures that are policy-relevant in their respective countries. Second, they employed the EQ-5D-3L for evaluation. These factors significantly influenced the design of this study in a Japanese setting. Second, regarding the choice of OAB measure, we used the OABSS, similar to Kim *et al*., as it is explicitly used to define OAB in the Japanese OAB guidelines [23]. The OABSS was used to define OAB in a randomized controlled trial evaluating the effects of OAB interventions in Japan [24]. Therefore, decision modelling based on the OABSS is expected when conducting HEEs targeting patients with OAB in Japan. Second, we used the EQ-5D-5L, for which a value set developed and validated by Ikeda *et al*. exists in Japan [17]. Evidence suggests that the EQ-5D-3L may potentially overestimate positive associations and underestimate negative associations compared with the EQ-5D-5L in HEEs [15]. Consequently, the EQ-5D-5L is recommended for cost-effectiveness evaluations in Japan [25], and our study adhered to this recommendation. This study, which fully considers Japanese clinical practices and healthcare systems, provides HUVs appropriate for HEEs in the Japanese setting. We believe that our approach offers valuable insights for the design of future studies in any country.

In this study, the disutility values for mild and moderate-to-severe OABs as defined by the OABSS were –0.034 and –0.0591, respectively. When referring to the Japanese population norms using the EQ-5D-5L, these disutilities are comparable to those of other chronic conditions that affect HRQoL, such as allergic rhinitis (–0.027), atopic dermatitis (–0.031), and other skin diseases (–0.043) [26]. In a report from Korea, which used the same scale (OABSS) and criteria to define OAB as our study, the covariate-adjusted disutility values for mild OAB, moderate OAB, and severe OAB were –0.029, –0.089, and –0.179, respectively [14]. The smaller disutility for moderate-to-severe OAB in our study compared to Kim *et al*.’s findings may be explained by differences in the mean age (75 years vs. 45.2 years), outcome measure (EQ-5D-5L vs. EQ-5D-3L), and ethnicity (Japanese vs. Korean). However, both studies consistently showed that disutility increased with severity, emphasizing the importance of the associations of OAB with HUVs. Furthermore, our study found high disutility (–0.1026) among female participants with moderate-to-severe OAB in the subgroup analyses. This finding may be supported by the results of EpiLUTS, an Internet survey conducted among individuals aged >40 years in the United States, which showed that a higher proportion of women (39.4%) than men (28.7%) reported moderate-to-severe problems in their patient perception of bladder condition for OAB with bother [27]. Additionally, in the subgroup of individuals aged ≥75 years, mild and moderate-to-severe OABs were associated with disutility values of similar magnitude (mild OAB –0.0688 vs. moderate-to-severe OAB –0.0664). This finding suggests that the presence of OAB, regardless of severity, may have a substantial impact on older adults who are more likely to be frail than younger respondents. For example, evidence has shown that even in the absence of incontinence, the presence of OAB is a significant risk factor for falls [28].

Our study has several strengths. First, we comprehensively provided the descriptive and analytical information necessary for the HEE of OAB, specifically for older adults. The mean and median HUVs at baseline and the disutilities after covariate adjustment for each severity level of OAB are essential information when conducting HEEs. Second, this study estimated disutility by adjusting for a wide range of evidence-based covariates, from socioeconomic to health-related indicators. Third, a subgroup analysis was performed to identify vulnerable populations that are essential for HEEs from the perspective of equity considerations but are understudied [29]. Moreover, our subgroup analysis results provide HUVs that can be referenced in future HEEs, particularly when treatments specifically targeting nocturia or urgent urinary incontinence among the OAB symptoms become available.

However, this study has also some limitations. First, as this study utilized an Internet survey, there were concerns regarding the external validity of the results. However, the prevalence of OAB (15.9%) and mean HUV (0.89) in this study did not substantially differ from previous epidemiological data from Japan [26, 30]. Thus, these values are also expected to apply to HEEs. Second, the sample size was limited (approximately 1,000); consequently, there were few individuals with high scores on the OABSS (*e.g.* 2 points for daytime frequency and 4–5 points for urgency urinary incontinence). As a result, it was not possible to differentiate between moderate and severe OAB, and some scores were grouped in the exploratory analysis. Although the number of individuals with such high scores may not be large and may not be considered in actual HEEs, revalidation of our study with a larger sample size would be valuable. Third, there may have been residual confounding factors. For example, orthopedic factors, such as falls and fractures, can be considered in future models.

In conclusion, this study examined HUVs in older Japanese adults with and without OAB. The results also demonstrated that increasing the severity of OAB indicated greater disutility, highlighting the substantial effect of OAB on HRQoL. These findings serve as valuable resources for future HEEs of OAB, possibly guiding decision-making in healthcare resource allocation for policymakers.

## Data Availability

The datasets generated and analyzed during the current study are available from the corresponding author upon reasonable request.

## Acknowledgments

This study was supported by the Japan Society for the Promotion of Science KAKENHI grant (grant number: 21K17228) for conducting the survey. This study was also supported by the National Institute of Public Health for the language editing fee and article publishing charge.

