## Supplementary materials for "Health Utility Value of Overactive Bladder in Japanese Older Adults"

**Method S1** The questionnaires and response options in the survey

 What is your age?
 What is your sex?
  1. Male
  2. Female
  3. Others
 Please provide your current height and weight.
  1. Height (cm):
  2. Weight (kg):
 What is your annual household income? Please choose one of the following options:

^*^This includes the total income earned by your entire household in 1 year, including income from work and other sources (such as remittances from children, property income like rent, benefits for older adults).

1. Less than 1 million yen
2. 1 million yen to <2 million yen
3. 2 million yen to <4 million yen
4. 4 million yen to <6 million yen
5. 6 million yen to <10 million yen
6. 10 million yen to <15 million yen
7. 15 million yen to <20 million yen
8. ≥20 million yen
9. Prefer not to answer
10. Do not know
 Including yourself, how many family members share your household finances?
 Do you currently have a spouse (husband or wife, including a partner, in a de facto marital relationship)?
  1. No (I am single)
  2. Yes
  3. I had one, but they passed away (widowed).
  4. I had one, but we divorced.
 Do you currently smoke tobacco habitually?
  1. I smoke almost always.
  2. I smoke occasionally.
  3. I used to smoke in the past, but I do not smoke now.
  4. I do not smoke and have never smoked.
 Do you currently drink alcohol habitually?
  1. I drink almost always.
  2. I drink occasionally.
  3. I used to drink in the past, but I do not drink now.
  4. I do not drink and have never drunk.
 Have you experienced any of the following conditions? For each condition, we selected one of the following options:
  1. Never had it
  2. Had (or have) it, but not currently receiving treatment at a hospital
  3. Have it and currently receiving treatment at a hospital
  4. Prefer not to answer

Conditions:

1. Hypertension (high blood pressure)
2. Diabetes
3. Hyperlipidemia/hypercholesterolemia
4. Lower back pain/lumbago
5. Angina pectoris/myocardial infarction
6. Cerebral infarction/cerebral hemorrhage
7. Kidney disease/renal failure
8. Stress urinary incontinence (involuntary urination when coughing, sneezing, etc.)
9. Depression
10. Mental health conditions other than depression (requiring psychiatric or psychosomatic medicine)

OABSS

 How many times do you typically urinate from waking in the morning until sleeping at night?
  1. ≤7
  2. 8–14
  3. ≥15
 How many times do you typically wake up to urinate from sleeping at night until waking in the morning?
1. 0
2. 1
3. 2
4. ≥3
 How often do you have a sudden desire to urinate, which is difficult to defer?
  1. Not at all
  2. Less than once a week
  3. Once a week or more
  4. About once a day
  5. 4 times a day
  6. 5 times a day or more
 How often do you leak urine because you cannot defer the sudden desire to urinate?
  1. Not at all
  2. Less than once a week
  3. Once a week or more
  4. About once a day
  5. 4 times a day
  6. 5 times a day or more

In addition to the above questions, we used the Japanese versions of the EQ-5D-5L and EQ VAS. For information on the EQ-5D-5L and EQ VAS, please refer to the user guide available on the EuroQol website (https://euroqol. org/information-and-support/documentation/user-guides).

**Table S1** Baseline characteristics of the sex subgroups

| **Variables** | **Category** | **Male (n=516)** | | **Female (n=478)** | |
| --- | --- | --- | --- | --- | --- |
|  |  | **n** | **%** | **n** | **%** |
| Age | Mean (SD) | 73.3 | (5.3) | 73.1 | (5.3) |
|  | Median [IQR] | 75 | [68.5–77] | 74 | [68–76] |
| BMI | Mean (SD) | 23.1 | (2.8) | 21.9 | (3.8) |
|  | Median [IQR] | 22.9 | [21.3–23.7] | 21.6 | [19.7–23.7] |
| Educational attainment | College graduate or more | 322 | 62.4 | 235 | 49.2 |
| Equivalent household income | Lower half | 220 | 42.6 | 189 | 39.5 |
|  | Upper half | 201 | 39.0 | 159 | 33.3 |
|  | Decline to answer | 95 | 18.4 | 130 | 27.2 |
| Smoking status | Never | 97 | 18.8 | 380 | 79.5 |
|  | Past | 341 | 66.1 | 69 | 14.4 |
|  | Current | 78 | 15.1 | 29 | 6.1 |
| Alcohol consumption | Never | 52 | 10.1 | 194 | 40.6 |
|  | Past | 126 | 24.4 | 103 | 21.6 |
|  | Current | 338 | 65.5 | 181 | 37.9 |
| Presence of comorbidities | Hypertension | 288 | 55.8 | 198 | 41.4 |
|  | Dyslipidemia | 196 | 38.0 | 211 | 44.1 |
|  | Diabetes | 130 | 25.2 | 36 | 7.5 |
|  | Low back pain | 316 | 61.2 | 250 | 52.3 |
|  | Ischemic heart disease | 64 | 12.4 | 27 | 5.7 |
|  | Stroke | 48 | 9.3 | 15 | 3.1 |
|  | Chronic kidney disease | 21 | 4.1 | 15 | 3.1 |
|  | Stress urinary incontinence | 22 | 4.3 | 92 | 19.3 |
|  | Depression | 30 | 5.8 | 20 | 4.2 |
|  | Other psychiatric disorders | 21 | 4.1 | 23 | 4.8 |
| OABSS | Mean (SD) | 3.1 | (2.3) | 2.2 | (1.9) |
|  | Median [IQR] | 3 | [1–4] | 2 | [1–3] |
| HUV | Mean (SD) | 0.91 | (0.11) | 0.90 | (0.12) |
|  | Median [IQR] | 0.89 | [0.84–1] | 0.89 | [0.83–1] |

**Note.** BMI, body mass index; HUV, health utility value; IQR, interquartile range; OABSS, Overactive Bladder Symptom Scale; SD, standard deviation.

**Table S2** Baseline characteristics of the age subgroups

| **Variables** | **Category** | **65 to 74 (n=492)** | | **75 to 94 (n=502)** | |
| --- | --- | --- | --- | --- | --- |
|  |  | **n** | **%** | **n** | **%** |
| Age | Mean (SD) | 68.7 | (2.9) | 77.7 | (2.9) |
|  | Median [IQR] | 68 | [66–71] | 77 | [75–79] |
| Sex | Male | 250 | 50.8 | 266 | 53.0 |
| BMI | Mean (SD) | 22.5 | (3.2) | 22.6 | (3.5) |
|  | Median [IQR] | 22.5 | [20.3–24.3] | 22.2 | [20.8–24.3] |
| Educational attainment | College graduate or more | 309 | 62.8 | 248 | 49.4 |
| Equivalent household income | Lower half | 183 | 37.2 | 226 | 45.0 |
|  | Upper half | 184 | 37.4 | 176 | 35.1 |
|  | Decline to answer | 125 | 25.4 | 100 | 19.9 |
| Smoking status | Never | 229 | 46.5 | 248 | 49.4 |
|  | Past | 197 | 40.0 | 213 | 42.4 |
|  | Current | 66 | 13.4 | 41 | 8.2 |
| Alcohol consumption | Never | 115 | 23.4 | 131 | 26.1 |
|  | Past | 116 | 23.6 | 113 | 22.5 |
|  | Current | 261 | 53.1 | 258 | 51.4 |
| Presence of comorbidities | Hypertension | 214 | 43.5 | 272 | 54.2 |
|  | Dyslipidemia | 203 | 41.3 | 204 | 40.6 |
|  | Diabetes | 83 | 16.9 | 83 | 16.5 |
|  | Low back pain | 273 | 55.5 | 293 | 58.4 |
|  | Ischemic heart disease | 39 | 7.9 | 52 | 10.4 |
|  | Stroke | 28 | 5.7 | 35 | 7.0 |
|  | Chronic kidney disease | 13 | 2.6 | 23 | 4.6 |
|  | Stress urinary incontinence | 52 | 10.6 | 62 | 12.4 |
|  | Depression | 30 | 6.1 | 20 | 4.0 |
|  | Other psychiatric disorders | 25 | 5.1 | 19 | 3.8 |
| OABSS | Mean (SD) | 2.4 | (2.0) | 2.9 | (2.3) |
|  | Median [IQR] | 2 | [1–3] | 2 | [1–4] |
| HUV | Mean (SD) | 0.92 | (0.10) | 0.89 | (0.12) |
|  | Median [IQR] | 0.89 | [0.87–1] | 0.89 | [0.83–1] |

**Note.** BMI, body mass index; HUV, health utility value; IQR, interquartile range; OABSS, Overactive Bladder Symptom Scale; SD, standard deviation.

**Table S3** Prevalence of OAB and utility values according to sex or age subgroups

| **Population** | **OAB severity^*^** | **n (%)** | **Mean HUV (SD)** | **Median HUV [IQR]** |
| --- | --- | --- | --- | --- |
| **Sex subgroups** |  |  |  |  |
| **Male (n=516)** | **Non-OAB** | **409 (79.3)** | **0.917 (0.104)** | **0.895 [0.875–1]** |
|  | **Any OAB** | **107 (20.7)** | **0.872 (0.128)** | **0.895 [0.823–1]** |
|  | Mild OAB | 43 (8.3) | 0.882 (0.119) | 0.895 [0.823–1] |
|  | Moderate-to-severe OAB | 64 (12.4) | 0.865 (0.134) | 0.895 [0.811–1] |
| **Female (n=478)** | **Non-OAB** | **427 (89.3)** | **0.909 (0.107)** | **0.894 [0.867–1]** |
|  | **Any OAB** | **51 (10.7)** | **0.821 (0.173)** | **0.831 [0.759–0.895]** |
|  | Mild OAB | 28 (5.9) | 0.862 (0.150) | 0.895 [0.824–1] |
|  | Moderate-to-severe OAB | 23 (4.8) | 0.770 (0.189) | 0.823 [0.670–0.895] |
| **Age subgroups** |  |  |  |  |
| **65–74 years (n=492)** | **Non-OAB** | **423 (86.0)** | **0.920 (0.100)** | **0.895 [0.875–1]** |
|  | **Any OAB** | **69 (14.0)** | **0.889 (0.130)** | **0.895 [0.823–1]** |
|  | Mild OAB | 37 (7.5) | 0.916 (0.091) | 0.895 [0.829–1] |
|  | Moderate-to-severe OAB | 32 (6.5) | 0.858 (0.160) | 0.895 [0.766–1] |
| **75–94 years (n=502)** | **Non-OAB** | **413 (82.3)** | **0.907 (0.111)** | **0.894 [0.867–1]** |
|  | **Any OAB** | **89 (17.7)** | **0.829 (0.152)** | **0.831 [0.776–0.895]** |
|  | Mild OAB | 34 (6.8) | 0.829 (0.153) | 0.849 [0.780–0.895] |
|  | Moderate-to-severe OAB | 55 (11.0) | 0.830 (0.152) | 0.831 [0.759–1] |

**Note.** HUV, health utility value; IQR, interquartile range; OAB, overactive bladder; SD, standard deviation.

^*^Mild OAB is defined as domain 3 (urgency) of overactive bladder symptom score (OABSS)≥2 and total score of 3≤OABSS≤5, and moderate-to-severe OAB is defined as domain 3 of OABSS≥2 and total score of 6≤OABSS≤15.

**Table S4** Proportion and utility values of each score according to the OABSS domains

| **Domain** | **Score** | **n (%)** | **Mean HUV (SD)** | **Median HUV [IQR]** |
| --- | --- | --- | --- | --- |
| **Daytime frequency** | 0 | 549 (55.2) | 0.905 (0.113) | 0.895 [0.831–1] |
|  | 1 | 430 (43.3) | 0.905 (0.114) | 0.895 [0.844–1] |
|  | 2 | 15 (1.5) | 0.854 (0.158) | 0.895 [0.759–1] |
| **Nighttime frequency** | 0 | 195 (19.6) | 0.924 (0.096) | 1 [0.895–1] |
|  | 1 | 463 (46.6) | 0.909 (0.106) | 0.895 [0.867–1] |
|  | 2 | 228 (22.9) | 0.894 (0.116) | 0.895 [0.831–1] |
|  | 3 | 108 (10.9) | 0.869 (0.160) | 0.895 [0.823–1] |
| **Urgency** | 0 | 557 (56.0) | 0.920 (0.096) | 0.895 [0.875–1] |
|  | 1 | 279 (28.1) | 0.899 (0.121) | 0.895 [0.823–1] |
|  | 2 | 88 (8.9) | 0.866 (0.150) | 0.895 [0.823–1] |
|  | 3 | 42 (4.2) | 0.843 (0.141) | 0.873 [0.772–1] |
|  | 4 | 22 (2.2) | 0.842 (0.148) | 0.856 [0.759–1] |
|  | 5 | 6 (0.6) | 0.825 (0.117) | 0.831 [0.708–0.895] |
| **Urgency incontinence** | 0 | 818 (82.3) | 0.910 (0.108) | 0.895 [0.867–1] |
|  | 1 | 132 (13.3) | 0.896 (0.120) | 0.895 [0.831–1] |
|  | 2 | 27 (2.7) | 0.804 (0.175) | 0.831 [0.708–0.895] |
|  | 3 | 12 (1.2) | 0.842 (0.139) | 0.881 [0.741–0.947] |
|  | 4 | 5 (0.5) | 0.753 (0.185) | 0.759 [0.641–0.844] |
|  | 5 | 0 (0) | 0 (0) | 0 [0–0] |

**Note.** HUV, health utility value; IQR, interquartile range; OABSS, Overactive Bladder Symptom Score; SD, standard deviation.
